# "Stroke Patient and Stakeholder Engagement (SPSE): Concepts, Definitions, Models, Implementation Strategies, Indicators, and Frameworks - A Systematic Scoping Review"

**DOI:** 10.1101/2024.07.03.24309878

**Authors:** Hamidreza Khankeh, Gordon Guyatt, Shima Shirozhan, Juliet Roudini, Torsten Rackoll, Ulrich Dirnagl

**Author notes:** **Corresponding author:** Ulrich Dirnagl.

## Abstract

**Background:** Involving stroke patients in clinical research through patient engagement aims to ensure that studies are patient-centered, and may help ensure they are feasible, ethical, and credible, ultimately leading to enhanced trust and communication between researchers and the patient community. In this study, we have conducted a scoping review to identify existing evidence and gaps in SPSE.

**Methods:** The five-step approach outlined by Arksey and O’Malley, in conjunction with the Preferred Reporting Items for Scoping Reviews (PRISMA-ScR) guidelines, provided the structure for this review. To find relevant articles, we searched PubMed, Web of Science, and Embase databases up to February 2024. Additionally, the review team conducted a hand search using Google Scholar, key journals, and references of highly relevant articles. Reviewers conducted primary and secondary screening, ultimately selecting English-language articles with available full texts that met the eligibility criteria. Reviewers extracted data from these articles into a table designed and tested by the research team.

**Results:** Of 1,002 articles initially identified, 21 proved eligible. Stakeholder engagement primarily occurred during the design phase of studies and within the studies using qualitative methodologies. Although the engagement of stakeholders in the research process is increasing, practice regarding terminology and principles of implementation remains variable. Researchers have recognized the benefits of stakeholder engagement, but have also faced numerous challenges that often arise during the research process.

**Conclusion:** The current study identifies stakeholder groups and the benefits and challenges researchers face in implementing their engagement. Given existing challenges and limited specific models or frameworks, it is recommended to explore applied recommendations for stakeholder engagement in future studies, that may enhance stakeholder engagement, overcome obstacles, and unify researchers’ understanding of engagement and implementation.

## Introduction

Involving stroke patients in clinical research through Patient and Stakeholder Engagement (PSE), also known as Patient and Public Involvement (PPI), helps ensure that studies are patient-centered, and can enhance feasibility, credibility, and ethical conduct (1–4). This approach can ultimately lead to more relevant and effective outcomes, as well as improved trust and communication between researchers and the patient community. Additionally, it is expected to enhance the overall quality of studies (5–8).

High-level organizations (e.g., UNESCO, EU) and well-known funders (Bill and Melinda Gates Foundation, Wellcome Trust, etc.) are increasingly mandating PSE for the clinical studies they fund. Despite the numerous papers addressing PSE activities and various recommendations of differing quality and focus, there remains a lack of consensus among authors regarding the most effective methods for developing and nurturing PSE (4, 6, 9–11). There is no well-established framework or model, the terminology is often unclear, and ultimately, more evidence is needed to define the conditions and approaches under which PSE is most effective (6, 9–11).The unclear definitions and diverse terminology can exacerbate the special challenges that individuals with disabilities such as communication and cognitive impairments seen in stroke patients face in research engagement (4, 12–14).

Due to its status as the world’s second leading cause of death and long-term disability and its increasing frequency as populations age (15–17), we have therefore conducted a scoping review focusing on stroke as an index indication. Our ability to prevent and manage stroke, and thus decrease its burden, also influenced our choice. Optimal stroke prevention and management requires collaboration among governments, scientific organizations, healthcare professionals, researchers, patients, and families (15, 17–19). The European Stroke Action Plan (2018–2030) emphasizes improving the linkage between research results and patient populations (20). As in other indications, patient and stakeholder engagement in stroke research is not clear, necessitating comprehensive studies to identify barriers, gaps, needs, and opportunities regarding PSE. Although we focus on stroke, the findings will also be relevant to other diseases.

The primary objectives of this study are to gather evidence on Stroke Patient and Stakeholder Engagement (SPSE), derive insights from past experiences and recommendations, and clarify key concepts, definitions, models, strategies, indicators, and frameworks for establishing SPSE. Based on our findings we propose a comprehensive framework that integrates patient and stakeholder perspectives into stroke research, potentially advancing both theoretical understanding and practical applications for more effective, inclusive, and patient-centered stroke management and rehabilitation.”

## Methods

The study protocol entitled “Systematic scoping review protocol of Stroke Patient and Stakeholder Engagement (SPSE)” was published in 2023, in the Journal of Systematic Reviews (1). The planning, conducting, and reporting of the findings of this scoping review were guided by 5 steps described by Arksey and O’Malley -(1) identifying the initial research question, (2) identifying relevant studies, (3) study selection, (4) charting the data, and (5) collecting, summarizing, and reporting the results-as well as the Preferred Reporting Items for Scoping reviews (PRISMA-ScR) (21–23).

Systematic scoping reviews are fundamentally undertaken to map certain knowledge fields, identify key concepts and knowledge gaps, as well as address thorough inquiries, which may involve a variety of approaches but do not evaluate the quality of studies (21–23).

- Step 1: Identifying the initial research question

A scoping review was employed to achieve the following objectives:

(1) Identifying the sorts of current SPSE evidence, models, or strategies for establishing SPSE;
(2) Clarifying the main concepts, definitions, and components of SPSE;
(3) Compiling the experiences, prerequisites, or suggestions for adopting or applying SPSE.

We addressed the following questions concerning research related to stroke:

1. What are the key concepts, definitions, components, models, indicators, or frameworks for establishing an SPSE?
2. What engagement principles or strategies are reported in the literature that could be used to plan, conduct, and/or disseminate stroke research findings in partnership with research users?

The findings of this scoping review will be utilized to shape the development of principles, models, or tools that will offer guidance to researchers in the field of SPSE.

- Step 2: Identifying relevant studies

Based on past studies, there are no clear and uniform terms in the field of stakeholder engagement in the research process (12, 24). To find relevant studies, a brief review of the existing studies and MeSH and Emtree terms was conducted to select the best keywords to search. Then, the search strategy was set with keywords “research”, “Cerebrovascular Disorders”, “stroke”, “Patient Advocacy”, “Caregivers”, and “Stakeholder Participation”.

Three health databases including PubMed, Web of Sciences, Embase, and Google Scholar search engine were searched until February 2024 to find related literature. Also, to complete the review, a hand search of studies and a review of resources of highly relevant articles and key journals were done. You can find the search strategy of the PubMed database in Supplemental Table 1.

Results from the search strategies were exported and managed using Endnote X.7.5.3 and Microsoft Word. The de-duplication process was completed using the steps outlined by Bramer et al (25). Two team members [ShSh and HKh] independently used a Word screening tool and the abstract-level eligibility criteria to screen titles and abstracts.

Based on inclusion criteria, English articles related to stroke patients or rehabilitated patients with research competence in PSE are considered for reporting in this scoping review study.

- Step 3: Study selection

The final studies were selected after secondary screening by reading the full text of the articles, which was based on the inclusion and exclusion criteria. The inclusion and exclusion criteria are presented in Table 1. The full-text screening process was conducted independently by the same screening pairs. Consensus discussions were held to resolve any disagreements between screeners. If disagreements could not be resolved throughout the discussion, a third team member (JR or TR) was consulted to resolve them.

- Step 4: Charting Data and data extraction

**Table 1:**
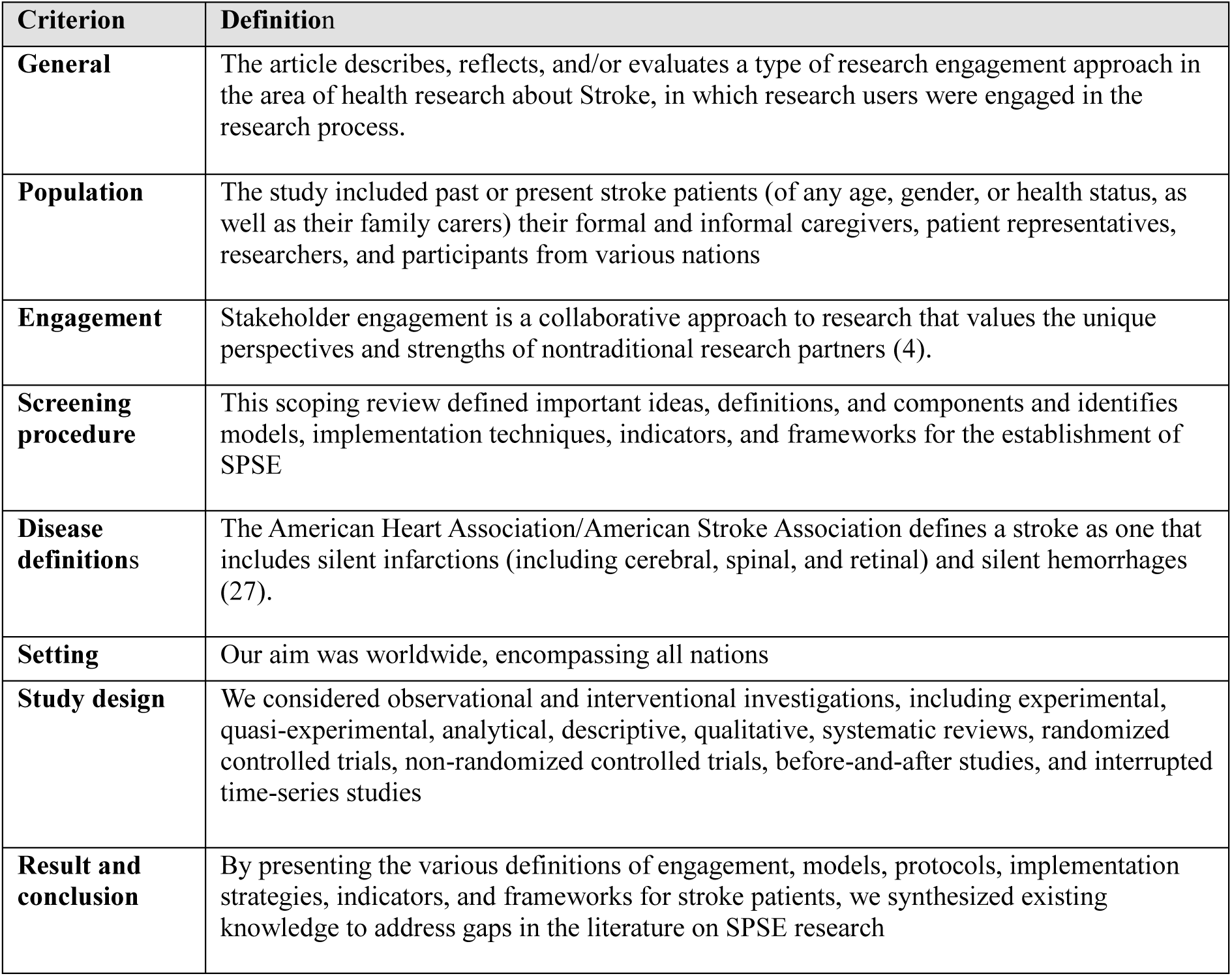
The inclusion and exclusion criteria of the study.

| **Criterion** | **Definition** |
| --- | --- |
| **General** | The article describes, reflects, and/or evaluates a type of research engagement approach in the area of health research about Stroke, in which research users were engaged in the research process. |
| **Population** | The study included past or present stroke patients (of any age, gender, or health status, as well as their family carers) their formal and informal caregivers, patient representatives, researchers, and participants from various nations |
| **Engagement** | Stakeholder engagement is a collaborative approach to research that values the unique perspectives and strengths of nontraditional research partners (4). |
| **Screening procedure** | This scoping review defined important ideas, definitions, and components and identifies models, implementation techniques, indicators, and frameworks for the establishment of SPSE |
| **Disease definitions** | The American Heart Association/American Stroke Association defines a stroke as one that includes silent infarctions (including cerebral, spinal, and retinal) and silent hemorrhages (27). |
| **Setting** | Our aim was worldwide, encompassing all nations |
| **Study design** | We considered observational and interventional investigations, including experimental, quasi-experimental, analytical, descriptive, qualitative, systematic reviews, randomized controlled trials, non-randomized controlled trials, before-and-after studies, and interrupted time-series studies |
| **Result and conclusion** | By presenting the various definitions of engagement, models, protocols, implementation strategies, indicators, and frameworks for stroke patients, we synthesized existing knowledge to address gaps in the literature on SPSE research |

For data extraction and presentation of the review results, the authors designed and tested several tables and figures, which can be found in the results section. Two researchers (ShSh and HKh) independently extracted the data and afterward integrated their findings. Data extraction and analysis followed the directed qualitative content analysis method suggested by Hsieh and Shannon, utilizing previously established definitions, principles, terminologies, and a comprehensive list of related information from published papers (26).

The study characteristics extracted utilizing an Excel form and exported to Word included the first author, year of publication, country of study, title, study design, and study aims. The engagement characteristics extracted were the definition of stakeholder engagement, group of engaged stakeholders, phase of stakeholder engagement, challenges and benefits of stakeholder engagement, and principles and strategies to implement SPSE in the research process.

The first author (HKh) reviewed the extracted information and resolved any uncertainties (e.g., engagement definitions or study design) through discussions with two team members (JR or TR). Subsequently, two team members (HKh and ShSh) who contributed to data extraction reviewed the findings and provided feedback. The first author (HKh) then finalized the results based on this feedback. Finally, each category of extracted data was referenced to the corresponding publications from which they were extracted.

- Step 5: Collecting, summarizing, and reporting the results

A summary and report of findings are provided below in the results part.

## Results

A systematic search resulted in 1,450 articles, of which 438 were removed due to duplication. In the first screening, the title and abstract of the remaining 1002 articles were reviewed. Based on inclusion and exclusion criteria, 63 articles were selected for full-text reviewing and secondary screening. Ultimately, 21 articles met the inclusion criteria and were reported in this scoping review study (Figure 1).

**Figure 1:**
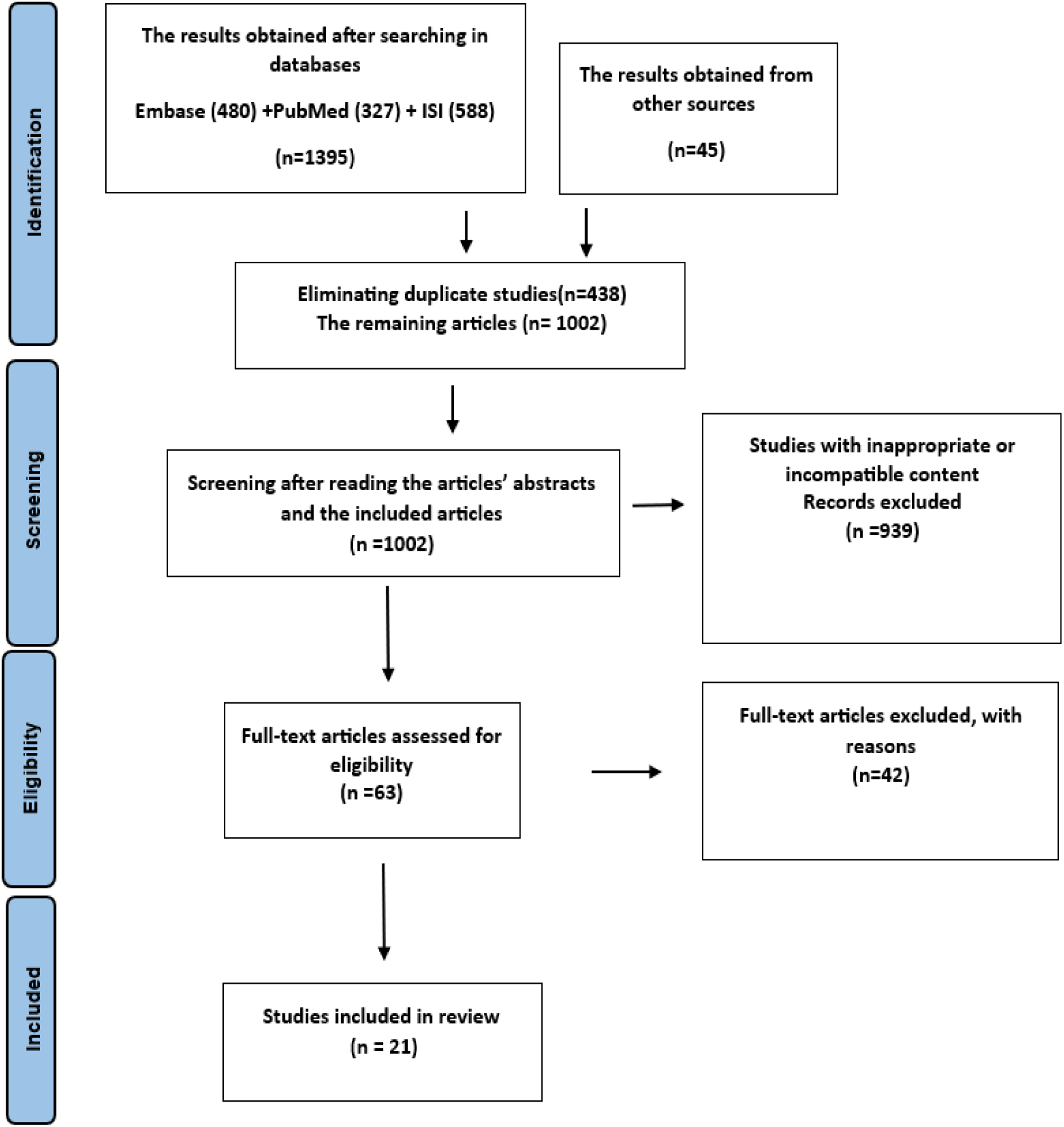
The PRISMA flowchart

European countries have been at the forefront of SPSE studies, with the UK leading with five papers, followed by the US and China with three each. Other studies have been conducted in Italy, Australia, Canada, Germany, Norway, Sierra Leone, India, Nigeria and Scotland (Figure 2).

**Figure 2:**
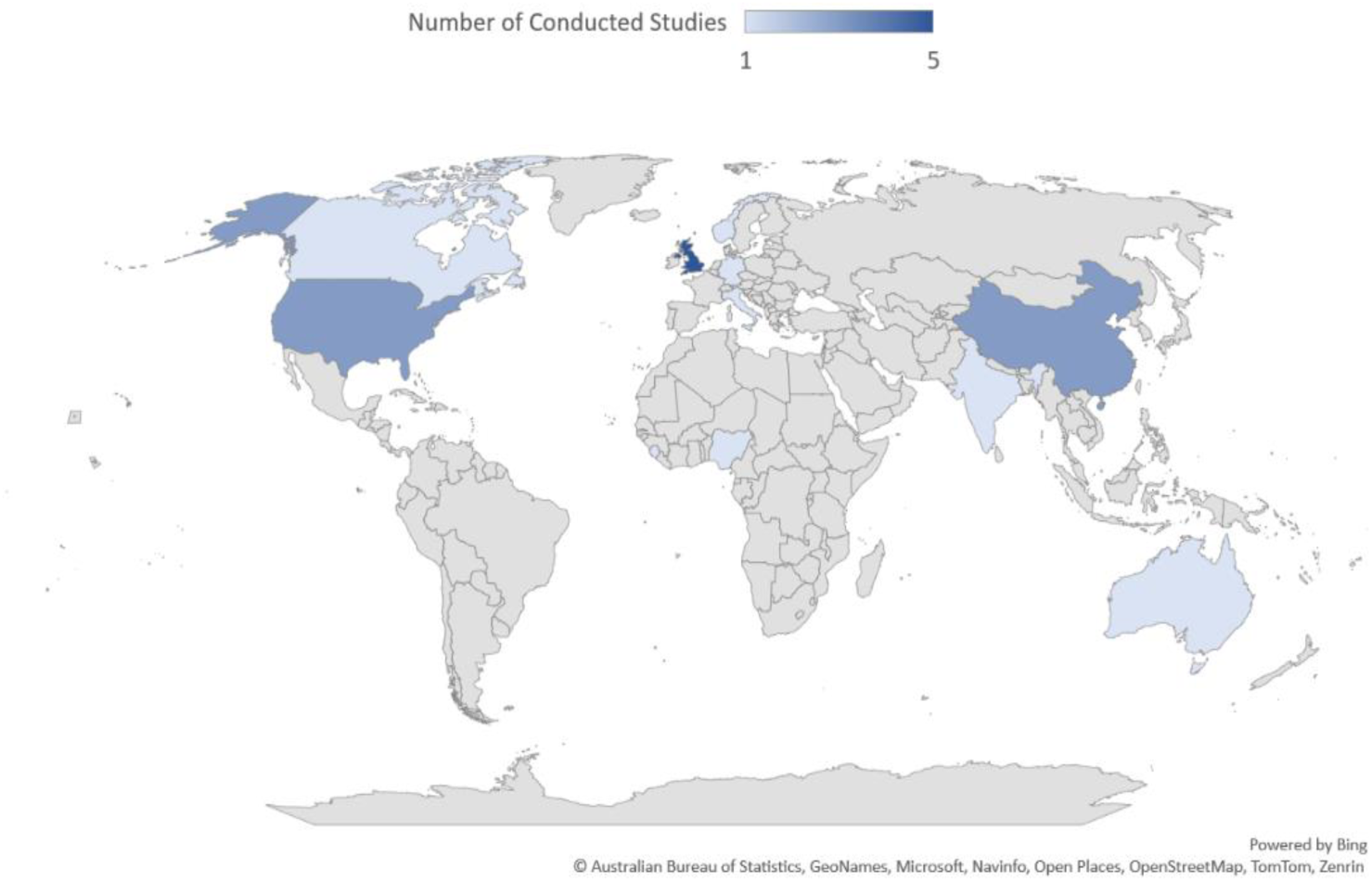
Spatial distribution of published studies

Studies on SPSE are categorized based on their goals. 43% of the articles specifically focus on SPSE (first category), while 57% of studies address aims such as identity reconstruction, stroke recovery, or rehabilitation, all involving stakeholders in the research process (second category).

Research on SPSE has garnered attention from researchers since 2005. The number of published articles has steadily increased, with 61% of articles published after 2021, averaging three studies per year (Figure 3). In recent years, the number of articles that have engaged stakeholders (second category) has proved greater than those that have discussed SPSE (first category). The number of articles published in 2024 may increase by the end of the year.

**Figure 3:**
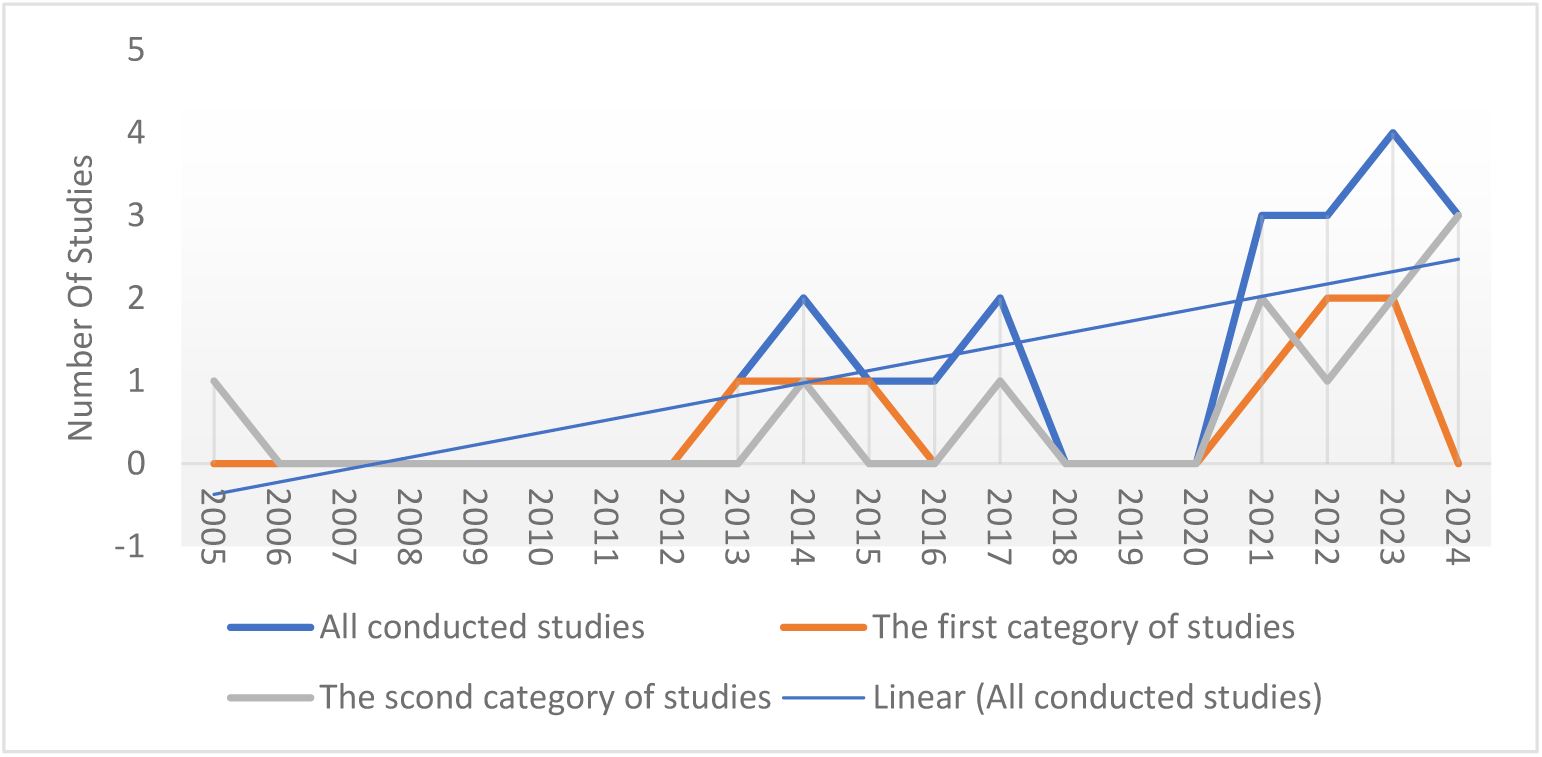
Temporal Trends in Published SPSE Studies

The first category of studies that specifically focused on SPSE is shown in Table 2. A significant number of studies did not have a clearly defined methodological approach. None of the studies that referenced the SPSE model presented a comprehensive and transparent outline of how stroke patients and their stakeholders could participate in the research. Additionally, none of the studies outlined any guidelines about engagement. Although the involvement of patients and stakeholders in different phases of the research was mentioned in 67% of the articles, less than half of the studies acknowledged the complications associated with engagement. Intriguingly, more than 77% of the studies proposed recommendations for enhancing SPSE, as shown in Table 2.

**Table 2:** Studies that specifically focused on SPSE (First category)

|  | Author(s) | Title | Type of study | Key findings |
| --- | --- | --- | --- | --- |

|  |  |  |  | Model | Guideline | Process of engagement | Challenges | Recommendations |
| --- | --- | --- | --- | --- | --- | --- | --- | --- |
| 1 | Hoekstra et al. (12) | Systematic overviews of partnership principles and strategies identified from health research about spinal cord injury and related health conditions: A scoping review | Scoping review |  |  | * |  | * |
| 2 | Brown et al. (14) | Stakeholder involvement in a Cochrane review of physical rehabilitation after stroke: Description and reflections | Not clear |  |  | * | * | * |
| 3 | Arnold et al. (28) | “What Do You Need? What Are You Experiencing?” Relationship Building and Power Dynamics in Participatory Research Projects: Critical Self-Reflections of Researchers | Not clear |  |  |  | * | * |
| 4 | McShan et al. (8) | Better Together: Evolution of Patient Stakeholder Engagement in Healthy Lifestyle Research After Acquired Brain Injury | Not clear |  |  | * |  |  |
| 5 | Roach et al. (29) | Lay Stakeholders in Science and Research Initiative | Not clear |  |  | * | * | * |
| 6 | Gesell et al. (4) | Methods guiding stakeholder engagement in planning a pragmatic study on changing stroke systems of care | Not clear | * |  | * |  | * |
| 7 | Boote et al. (30) | Stroke survivor and carer involvement in, and engagement with, studies adopted onto the NIHR Stroke Research Network portfolio: questionnaire survey | Letter to the editor |  |  | * |  |  |
| 8 | Pollock et al. (31) | Development of a new model to engage patients and clinicians in setting research priorities | priorities setting research | * |  |  |  | * |
| 9 | Sims et al. (32) | How to develop a patient and carer advisory group in stroke care research | Not clear |  |  |  | * | * |

The second category of articles includes studies that engage patients and stakeholders in the study process or have been planned for SPSE (study protocol). These protocols utilized a combination of qualitative (75%) and quantitative (25%) research methodologies. Various stakeholders, such as patients, caregivers/relatives, health professionals, and policymakers, were planned to be involved in the research process, including designing studies (2 articles), conducting studies (analyzing data, 1 article), and disseminating findings (1 article), as outlined in Table 3.

**Table 3:**
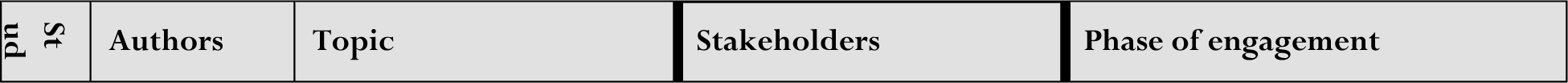
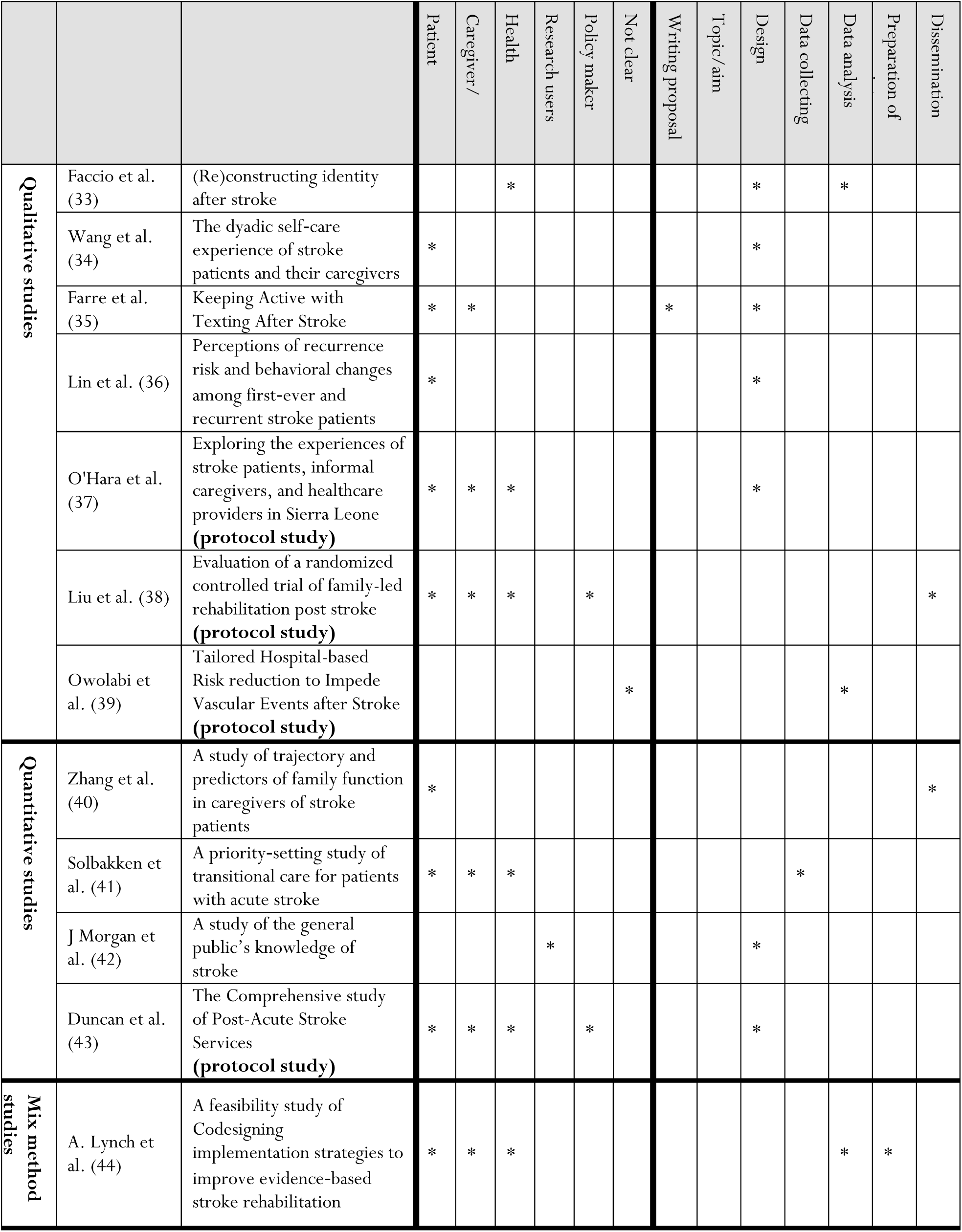
Studies that involve stakeholders in the research process (second category)

Completed studies have employed different research paradigms, including quantitative (3 articles), qualitative (4 articles), and mixed methods (1 article), to involve patients and stakeholders in the research process. These stakeholders included patients, caregivers/relatives, health professionals, and research users. One qualitative study specifically addressed the inclusion of SPSE in proposal writing. Engagement with stroke patients and their stakeholders occurred during the design phase in 62% of the articles, during data analysis in 25%, and in the dissemination of findings in 12% (Table 3).

The concept of Patient and Stakeholder Engagement (PSE) within the research process has been considered by various terms and phrases, including collaborative research approach, working together in a partnership, involvement of individuals who have an interest in the research, carrying out research with or by members of the public, an active partnership between patients, carers, and members of the public with researchers, engaging in collaborative research activities, the inclusion of users not as participants but as co-researchers, cooperation in decision-making and research activities which can be taken as co-production, codesigning, stakeholder involvement, and research partnerships (Table 4).

**Table 4:** Related terms/concepts to stakeholder engagement and definitions.

| Study | Stakeholder engagement and related concepts | Definition |
| --- | --- | --- |
| Gesell et al. (4) | Stakeholder Engagement | Stakeholder engagement is a collaborative research approach that values nontraditional research partners' unique perspectives and strengths. |
| Brown et al. (14) | Coproduction | Working together in a 'partnership' to produce research, and considered this one form of stakeholder involvement, in which specific criteria are met concerning partnership working |
| Brown et al. (14) | Stakeholder Involvement | The involvement in research of any people who have an interest in the research, in partnership with the (traditional) research team.<br>Research being carried out 'with' or 'by' members of the public rather than 'to,' 'about' or 'for' them. It is an active partnership between patients, carers, and members of the public with researchers that influences and shapes research |
| Hoekstra et al. (12) | Research Partnerships | Individuals, groups, or organizations that are engaged in collaborative research activities involving at least one researcher and any stakeholder. |
| Morgan et al. (42) | User Involvement | The inclusion of users not as participants but as co-researchers with a key position in the research design, data collection, interpretation, and dissemination processes. |
| Arnold et al. (28) | Participation | Participation refers to the cooperation in decision-making and research activities of those persons who are directly affected by the problems that are the subject of the respective research projects. |

Throughout phases of the research process, such as study design, implementation, and dissemination of results, diverse methodologies have been employed for engaging stakeholders, as outlined in Table 5. Among the 21 studies examined, only 6 (28%) provided detailed descriptions of the specific methods of stakeholder engagement utilized. These studies belong to the first category, which focuses on SPSE. Articles that engaged stakeholders in the research process did not elaborate on the methods of engagement.

**Table 5:** Different Stages of Patient and Stakeholder Engagement (PSE)

| Stage of study | Stakeholders' engagement | Resources |
| --- | --- | --- |
| Study design | ▪ Selection of the research topic<br>▪ Identifying or prioritizing topics for research<br>▪ Checking the applicability of the research<br>▪ Developing and revising the research question<br>▪ Elaboration or approval of the research proposal or protocol<br>▪ Compilation of the grant proposal<br>▪ Designing interventions | Boote et al. (30)<br>Gesell et al. (4)<br>Roach et al. (29)<br>McShan et al. (8)<br>Brown et al. (14)<br>Hoekstra et al. (12) |
|  | ▪ Design questions and interview guide<br>▪ Designing study inclusion criteria<br>▪ Develop, evaluate, or revise data collection tools<br>▪ Elaboration of informed consent form and details of participants |  |
| Study conduct | ▪ Literature review<br>▪ Selecting participants for the study<br>▪ Expressing opinions in various contexts such as forms of data collection and informed consent ‘study applications<br>▪ Conducting a pilot study and providing feedback<br>▪ Data collection<br>▪ Planning interviews<br>▪ Data analysis and interpretation<br>▪ Conducting interviews or managing focus group discussions<br>▪ Training participants in the field of interventions<br>▪ Supervision of study implementation | Boote et al. (30)<br>Gesell et al. (4)<br>Roach et al. (29)<br>McShan et al. (8)<br>Brown et al. (14)<br>Hoekstra et al. (12) |
| Dissemination of findings | ▪ Feedback on the comprehensibility and fluency of the published findings<br>▪ Feedback on the best strategies for disseminating findings (timeliness and effectiveness)<br>▪ Participating in preparing the manuscript of the article or reviewing and providing feedback<br>▪ Knowledge translation activities<br>▪ Translation of scientific data into comprehensive and understandable language<br>▪ Presentation of study results in conferences<br>▪ Providing clinical recommendations and policy briefs<br>▪ Planning on how to publish findings<br>▪ Developing key messages | Boote et al. (30)<br>Gesell et al. (4)<br>McShan et al. (8)<br>Brown et al. (14)<br>Hoekstra et al. (12) |

SPSE is considered from the early stages of research (e.g., topic/title selection) to the final stages (e.g., dissemination of findings). However, the majority of the articles did not provide comprehensive information on the mechanisms employed for engaging stakeholders in the planning, conducting, or communication of study findings.

The findings presented in Table 5 are applicable for conducting research within both quantitative and qualitative paradigms at different stages of the study. However, stakeholder engagement requires adherence to several principles, which can complicate the employment of SPSE at each stage of the research process. These principles are outlined in Table 6.

**Table 6:**
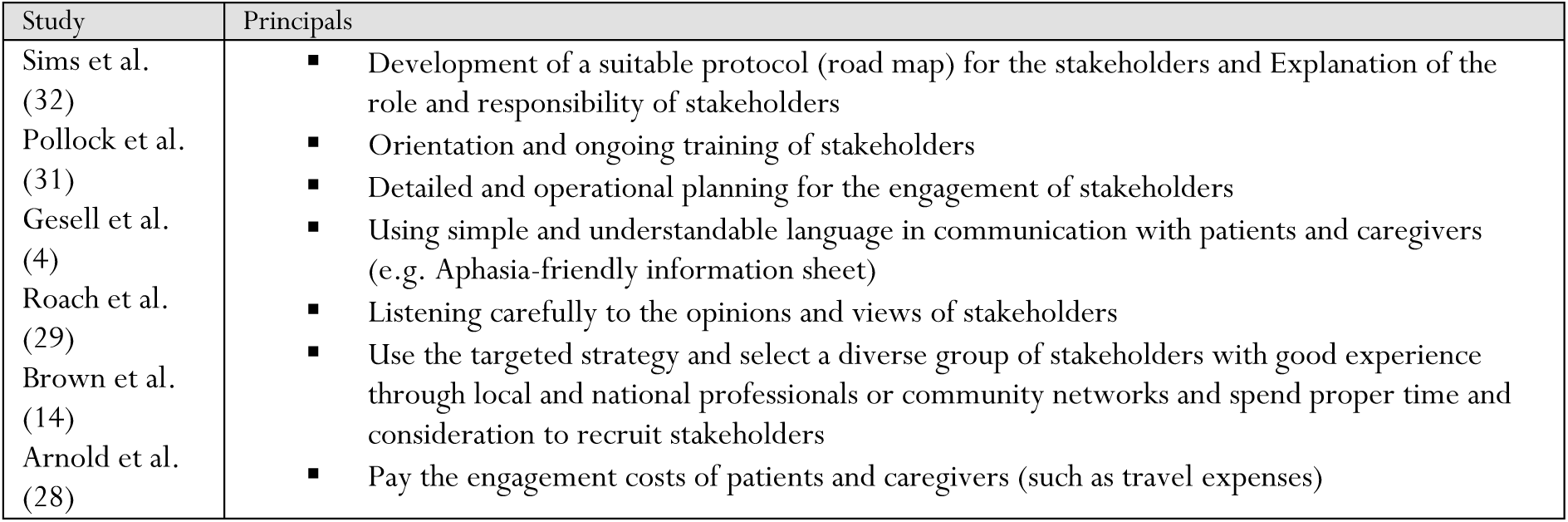
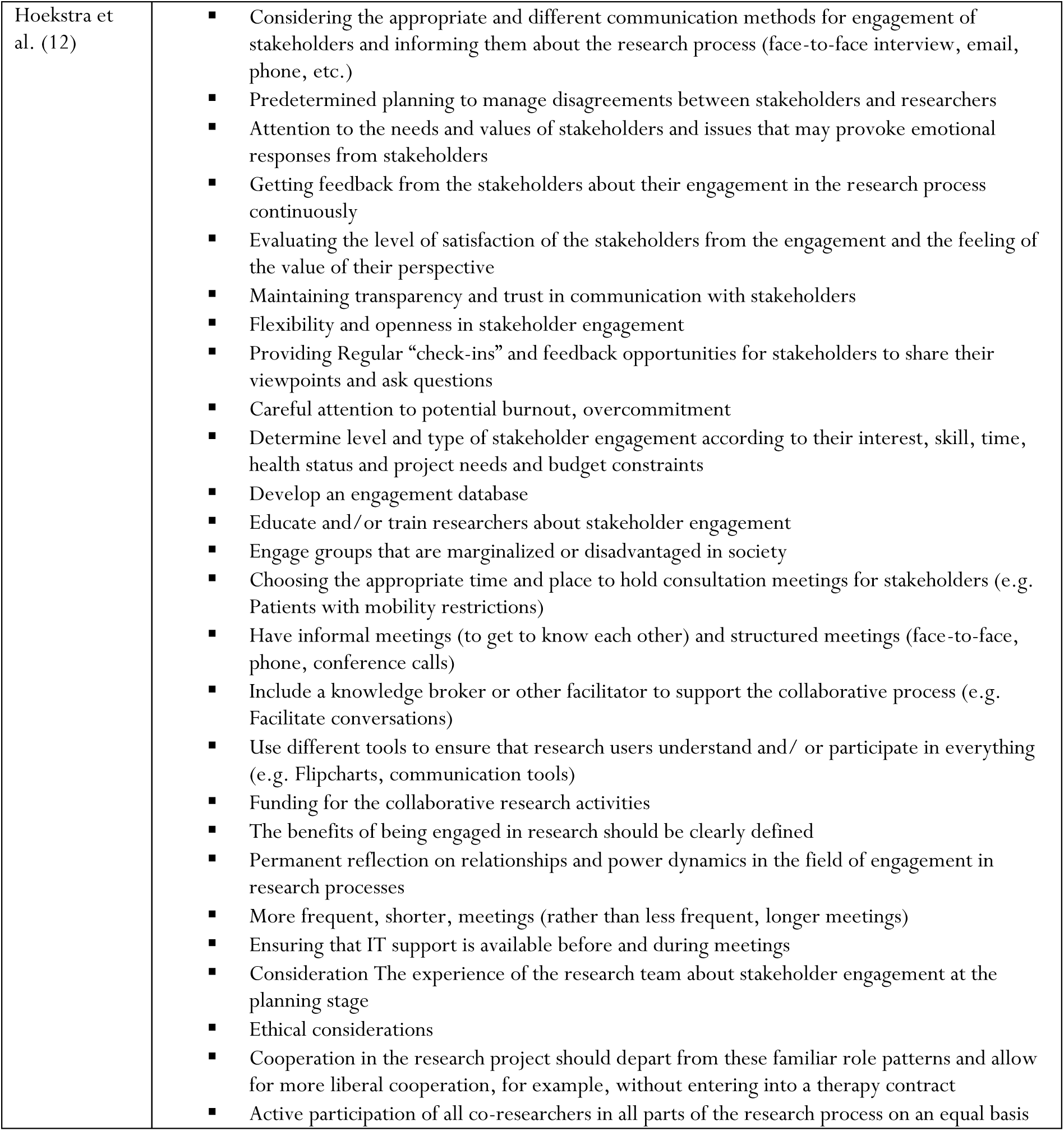
Principles of Patient and Stakeholder Engagement (PSE) in the Research Process.

This review highlights the limited integration of SPSE principles in previous studies, with only 33% of the articles reviewed demonstrating some aspects of SPSE. The analysis reveals a lack of consistency in the implementation of SPSE principles, with researchers often relying on their interpretations to guide their methodology of SPSE. The principles identified in the literature incorporate various aspects of the research process, including general principles (e.g., ethical consideration, flexibility, and openness) and specific guidelines, as summarized in Table 6.

Based on the findings, researchers should prepare for stakeholder engagement in multiple ways, such as developing a protocol or roadmap, allocating adequate resources in terms of cost and time, and providing continuous training to stakeholders. During the research process, it is crucial to ensure the active participation of stakeholders through measures such as soliciting feedback and holding informal meetings.

Articles discussing stakeholder engagement challenges accounted for only 14% of the total studies reported in Table 7. Our findings indicate that stakeholder engagement in the research process is associated with several challenges related to the research process, researchers, and stakeholders. Some challenges are inherent to the nature of engagement, such as being time-consuming and costly, which cannot be altered but can be managed. Other challenges, which pertain to all three areas, align with the principles mentioned in the previous table. Adhering to these principles, such as detailed and operational planning, appears to mitigate these challenges and can be further explored in future studies.

**Table 7:** Challenges regarding Patient and Stakeholder Engagement (PSE) in the research process.

| Related Area | Challenges | Resource |
| --- | --- | --- |
| Challenges related to stakeholders | Physical disabilities and communication disorders of patients (aphasia)<br>Fear of not being able to participate actively and being ineffective as a researcher<br>Feeling unable to balance their clinical responsibilities and duties in the research process | Sims et al. (32)<br>Arnold et al. (28) |
| Challenges related to researchers | Insufficient knowledge about how to engage stakeholders<br>Lack of enough time<br>Inadequate simplification and accessibility of stakeholder engagement in the research process | Sims et al. (32)<br>Brown et al. (14)<br>Arnold et al. (28) |
| Challenges related to the research process | The limitation of published studies that have experienced the engagement of stakeholders<br>Time-consuming and costly and limited resources for the engagement of stakeholders<br>Difference of opinions of researchers and stakeholders in the process of data analysis<br>Communication challenges between researchers and stakeholders (such as power relations)<br>Selection of the research topic<br>Determination of the level and stages of engagement<br>Organizational rules<br>Multiplicity of stakeholders | Sims et al. (32)<br>Gesell et al. (4)<br>Arnold et al. (28) |

Numerous articles have broadly examined the advantages of stakeholder engagement compared to studies focusing on challenges. Specifically, five studies have highlighted the benefits, while only three studies have addressed the challenges. Additionally, Table 8 illustrates that the anticipated benefits of stakeholder engagement outweigh the identified challenges. The proposed benefits of stakeholder engagement extend beyond stakeholders, researchers, and the research process, benefiting a wide range of groups.

**Table 8:**
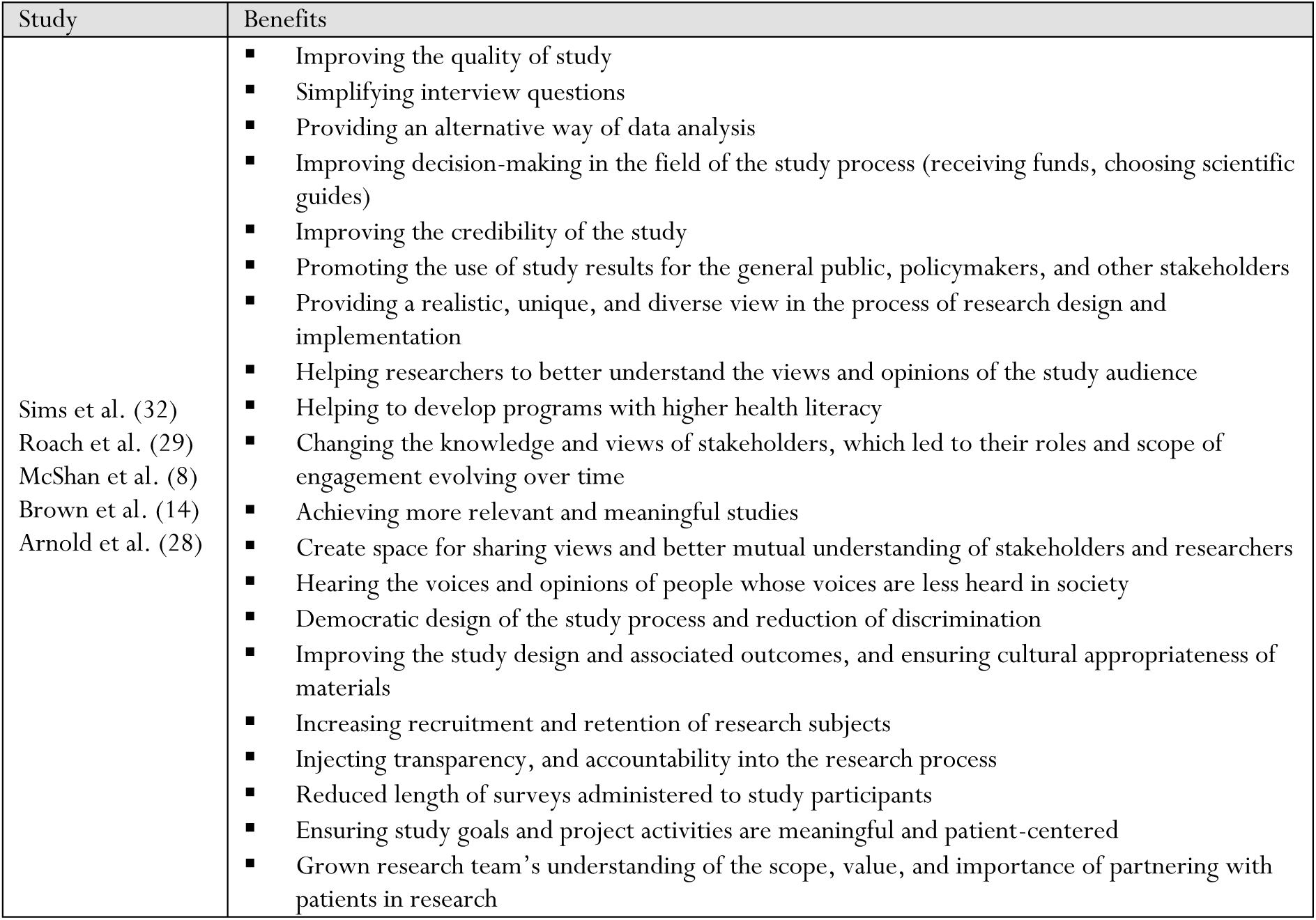
Benefits of Patient and Stakeholder Engagement (PSE) in the research process.

| Study | Benefits |
| --- | --- |
| Sims et al. (32)<br>Roach et al. (29)<br>McShan et al. (8)<br>Brown et al. (14)<br>Arnold et al. (28) | ▪ Improving the quality of study<br>▪ Simplifying interview questions<br>▪ Providing an alternative way of data analysis<br>▪ Improving decision-making in the field of the study process (receiving funds, choosing scientific guides)<br>▪ Improving the credibility of the study<br>▪ Promoting the use of study results for the general public, policymakers, and other stakeholders<br>▪ Providing a realistic, unique, and diverse view in the process of research design and implementation<br>▪ Helping researchers to better understand the views and opinions of the study audience<br>▪ Helping to develop programs with higher health literacy<br>▪ Changing the knowledge and views of stakeholders, which led to their roles and scope of engagement evolving over time<br>▪ Achieving more relevant and meaningful studies<br>▪ Create space for sharing views and better mutual understanding of stakeholders and researchers<br>▪ Hearing the voices and opinions of people whose voices are less heard in society<br>▪ Democratic design of the study process and reduction of discrimination<br>▪ Improving the study design and associated outcomes, and ensuring cultural appropriateness of materials<br>▪ Increasing recruitment and retention of research subjects<br>▪ Injecting transparency, and accountability into the research process<br>▪ Reduced length of surveys administered to study participants<br>▪ Ensuring study goals and project activities are meaningful and patient-centered<br>▪ Grown research team's understanding of the scope, value, and importance of partnering with patients in research |

## Discussion

This scoping review study identified types of evidence related to SPSE, summarized experiences and recommendations, clarified key concepts, definitions, and components, and identified models, implementation strategies, indicators, or frameworks for establishing SPSE.

The results of our study reveal a higher frequency of SPSE in qualitative research methodologies compared to quantitative and mixed methods which is in line with the study by Peniche, de Morais Faria et al. (45). Many of the studies reviewed lacked a defined methodological approach, and none provided a comprehensive and transparent framework for including stroke patients and their stakeholders in the research process. Furthermore, specific guidelines or protocols for engagement were not offered in any of the studies. While only a minority of the studies acknowledged the challenges related to engagement, most did provide general suggestions for SPSE. The majority of SPSE occurred during the design phase, followed by data analysis, and the dissemination of findings in completed studies.

SPSE has been described using various terms/concepts and phrases, all emphasizing a collaborative approach. Despite the importance of SPSE, the majority of articles reviewed did not carefully detail the strategies used to involve stakeholders in the research process. While a range of methodologies were utilized through the research process, only a limited number provided detailed descriptions of the SPSE methods employed. The literature defines principles of SPSE that cover various aspects of the research process, both general and specific, yet there is limited discussion on strategies specifically tailored to stroke patients and their stakeholders.

One of the strengths of our systematic scoping review is our comprehensive search and specific focus on stroke patients and their stakeholders, unlike previous reviews which had a more generalized approach to patient stakeholder engagements. Our use of clear eligibility criteria and rigorous methods for systematic data analysis and extraction ensures that we have not overlooked any essential information regarding characteristics, concepts, definitions, components, models, implementation strategies, indicators, challenges, benefits, or frameworks related to establishing stroke patient stakeholder engagements. We believe our study is comprehensive due to its inclusive exploration of recommendations and key concepts for establishing SPSE, distinguishing it from similar reviews. Our study involves all aspects of engagements, including PPI, without time constraints up to 2024. Following Arksey and O’Malley’s five steps and utilizing directed qualitative content analysis, we included all relevant papers, including protocols, categorizing them into those discussing or implementing engagement. Our study uniquely addresses the challenges and benefits related to researchers, stakeholders, and the research process in implementing SPSE.

This study was limited by the high number of conference papers and the restricted access to their full texts. Additionally, the diversity of terminology in the field of stakeholder engagement required the authors to formulate a search strategy based on MeSH Headings, Emtree terms, and synonyms. To address this limitation, hand searches were conducted to identify relevant studies and key journals, thereby planning to improve study accuracy. It is recommended that the terminology introduced in this study be integrated into the search strategies of future researchers.

Furthermore, the study was also constrained by the extensive studies utilizing co-design and priority-setting methods, which prevented their comprehensive inclusion in this scoping review. These methods warrant further investigation in future studies.

Based on this review, there has been a noticeable increase in the publication rate of articles, particularly since 2021 (8, 12–14, 28, 29, 33–37, 40, 41, 44). This trend aligns with a growing emphasis on stakeholder engagement in the research process, reflecting broader efforts towards knowledge translation, responsible research, and open science (46–48).

European countries, especially the United Kingdom have taken the lead in conducting SPSE studies. The increasing interest from other developed countries like the United States and China, as well as less developed countries such as Sierra Leone and Nigeria, suggests a growing interest in SPSE within the research community (4, 8, 34, 36, 37, 39, 40, 43). Despite concerns over the cost of SPSE, many suggested principles, such as prioritizing stakeholder needs and values, and maintaining flexibility and openness, appear to be achievable for researchers irrespective of financial constraints (14, 28, 32). Hence, it appears that implementing SPSE strategies is feasible across countries with varying economic and developmental statuses. Consequently, future research endeavors should focus on further exploring the advantages and impacts of engagement, including the economic and social repercussions of SPSE.

The findings of this review suggest that the articles can be categorized into two distinct groups based on the study’s objective; those that focused on discussing engagement (first category) and those that implemented this approach (second category) which is not considered in recently publish review (45).

The majority of the first group of studies that discussed SPSE lacked a defined methodological approach (4, 8, 14, 28, 29, 32). Additionally, none of the studies provided a comprehensive and transparent framework for including stroke patients and their stakeholders in the research process. Furthermore, none of the studies offered specific practical guidelines for engagement (4, 8, 12, 14, 28–32). Only a minority of the studies acknowledged the challenges related to engagement, but most of them did offer suggestions for SPSE (4, 12, 14, 28, 29, 31, 32).

The second group of studies on SPSE in Health Research had differing research paradigms, including qualitative, quantitative, or mixed methods, with some studies also following a protocol (33–36). Among these, articles with a qualitative paradigm were more commonly found. The authors of these studies suggest that the flexibility offered by qualitative research methods has contributed to the growing interest in stakeholder engagement in this area. Stakeholders have been involved in multiple stages of the research process, such as designing interview guides, collecting and analyzing data, and disseminating research findings (4, 8, 12, 14, 29, 30). However, challenges have been identified in the form of struggling perspectives and disagreements between researchers and stakeholders during the data analysis process (4, 14, 28, 32).

In quantitative research, stakeholders play a key role in the design and dissemination of research findings, as demonstrated by studies conducted by Zhang and Morgan (40, 42). Other studies also support the engagement of stakeholders in various stages of quantitative research, including selecting research topics and titles, formulating research questions, choosing appropriate measurement tools, designing interventions, and sharing research findings (40, 42–44).

Opposite to previous research in the field of SPSE, which primarily utilized quantitative and qualitative methodologies, only one mixed-methods study was identified. This study employed a co-design methodology (44).

In general, in the second category of studies, several stakeholders were involved in the research process, with the largest group being patients with stroke (37, 38, 43). Other stakeholders identified included caregivers and/or families of patients, professional teams, and policymakers. The terms ‘research users’ and ‘consumers’ were also used in Morgan and Hostetler’s study with a similar meaning of stakeholders, including a wide range of individuals within this group (12, 42). Wu et al.’s study on identifying stakeholders in healthcare introduced 12 groups as stakeholders in the healthcare system, including individuals from universities, hospitals, nursing homes, monitoring systems, and insurance institutions. The classification presented in this article appears to encompass all relevant people in healthcare and offers a comprehensive definition of stakeholders, which may be valuable for future researchers (49).

Regardless of the study paradigm in which the stakeholders engaged, the highest level of SPSE has been in the study design phase which is in line with the study by Peniche, de Morais Faria et al. followed by other stages including the formulation of a research proposal, data collection and analysis, drafting of the manuscript, and dissemination of research findings (33–36, 40, 41, 45).

Based on the results of this study, the terms "involvement" and "partnerships" have been identified as interchangeable with stakeholders’ engagement in the research process (4, 12, 14, 28, 42, 50). The use of various terms may present a challenge for researchers seeking to approach relevant studies on stakeholder engagement (24).Therefore, there is a serious need to establish a clear and comprehensive definition of stakeholder engagement in research to support future investigations (4, 24). Conducting concept analysis studies can help elucidate the nuances of this crucial concept and establish a comprehensive, widely accepted definition to facilitate coherence and consistency within the research community.

The methods of stakeholder engagement in the research process, from initial question formulation to dissemination of findings have been discussed in a few articles, which are in the first category. The dispersed and unstructured presentation of these steps in different studies makes it difficult to follow the SPSE step-by-step. Also, some steps are less discussed and remain unclear, further explanation of the process of engagement is needed (4, 8, 12, 14, 28–32).

In contrast to the inadequate methods mentioned for stakeholder engagement, several principles are considered in the articles. Key principles highlighted in these studies, such as ethical considerations, flexibility, and building trust, align with the guidelines for conducting research in the biomedical sciences. The unique characteristics of stroke patients and the diverse stakeholders involved necessitate the development of tailored protocols to effectively engage all parties involved (4, 14, 28, 29, 31, 32, 50).

However, given the diversity of research methodologies, the complex nature of the stroke patient’s complicated situation, and the numerous obstacles encountered during the research procedure, mere acquaintance with the principles and recommendations for designing and executing studies involving stakeholder engagement proves insufficient (4, 12, 51).

Also, the relationship between these principles and the methods of participation is not mentioned in the articles, and the prioritization of the implementation of these principles is not clear. It should also be noted that due to the different nature of studies in different paradigms, the importance and prioritization of these principles will be different (4, 12, 14, 28, 29, 31, 32).

Based on the findings of the study, challenges related to researchers, stakeholders, and the research process have been identified. Researchers may struggle with issues such as unfamiliarity with patient engagement and patients’ physical limitations (4, 14, 28, 32). Recommendations from various studies include allocating more resources and time for these types of studies, educating stakeholders, and increasing awareness before initiating research involving engagement (4, 12, 14, 28, 29, 31, 32). Differences between quantitative and qualitative paradigms, ethical concerns regarding stakeholders’ rights, potential compromises in study criteria, and the lack of evaluation standards for studies involving stakeholder engagement further complicate the issue (4, 8, 12, 14, 28–32).

In addition to the challenges associated with engaging stakeholders in the research process, several studies have highlighted the numerous advantages of this practice (46–48, 52). Many of these benefits, such as enhancing study quality, improving study credibility, and incorporating the perspectives of marginalized voices, align with principles of responsible research practices. The promotion of democracy, transparency, and patient-centeredness in research has garnered significant attention in recent years. The benefits of stakeholder engagement extend beyond just stakeholders, researchers, and the research process, benefiting various other groups of the community as well (46–48, 52). Further study is needed to explore the latent benefits of SPSE.

Based on the findings of existing studies, which indicate a lack of a specific framework or model, limited details on the steps and methods of stakeholder engagement, the need to observe several principles during the research process, and the presence of various challenges, it is evident that SPSE should be a focus of future research. Studies in this area should aim to provide a specific definition of stakeholder engagement, clarifying its antecedents, attributes, and consequences. Additionally, the development of specific models or frameworks and applied strategies for conducting studies in different paradigms should be followed.

## Conclusions

The research on Stroke Patient and stakeholder engagement (SPSE) has shown a global increase in recent years, with countries worldwide, including those in Europe, adopting this approach irrespective of their developmental stage. Although the reviewed literature often references related terms and definitions, there is a lack of substantial elaboration on them, indicating the need for more empirical studies to strengthen the concept and definitions. Stakeholder engagement has been detected across quantitative, qualitative, and mixed methods paradigms, predominantly occurring during the design phase. However, the research process using SPSE is often vague and controversial, underscoring the necessity for more qualitative studies to understand its nature and process. The existing principles in the literature cover different aspects related to research, offering effective guidance for studies in both quantitative and qualitative frameworks. Still, further research is required to explore hidden strategies and clarify their application. There is inconsistency in implementing SPSE principles, with researchers relying on personal interpretations. To enhance stakeholder engagement, researchers should plan for engagement diversely, allocate resources effectively, provide continuous training, and ensure active stakeholder participation. Challenges such as time and cost limitations are inherent in SPSE but can be managed with targeted strategies. Developing and implementing specific applied strategies tailored for SPSE can address challenges and pave the way for future research in this area. Establishing consistent terms and definitions for SPSE using a concept analysis study, can further advance research efforts.

## Data Availability

All data produced in the present work are contained in the manuscript

## List of abbreviations

PSE: Patient and Stakeholder Engagement
PPI: Patient and Public Involvement
SPSE: Stroke Patient and Stakeholder Engagement

## Declarations

### Ethics approval and consent to participate

Not applicable

### Availability of data and materials

The datasets used and/or analyzed during the current study are available from the corresponding author upon reasonable request.

### Authors’ contributions

HKh.: Conceptualization, Data curation, Investigation, Formal analysis, Funding acquisition, Methodology, Project administration, Writing – original draft, Writing - review & editing.

ShSh: Investigation, Data curation, Formal analysis, Writing - review & editing. JR.: Investigation, Writing - review & editing.

TR: Investigation, Writing - review & editing.

UD: Conceptualization, funding acquisition, methodology, Writing - review & editing, and supervision.

GG: Writing—review, and editing.

All authors gave final approval for publication and agreed to be held accountable for the work performed therein.

## Acknowledgements

We would like to thank Dr. Mohammad Saatchi for his support in writing study syntax to do a comprehensive search.

## Supplemental Files

**Supplemental Table 1:**
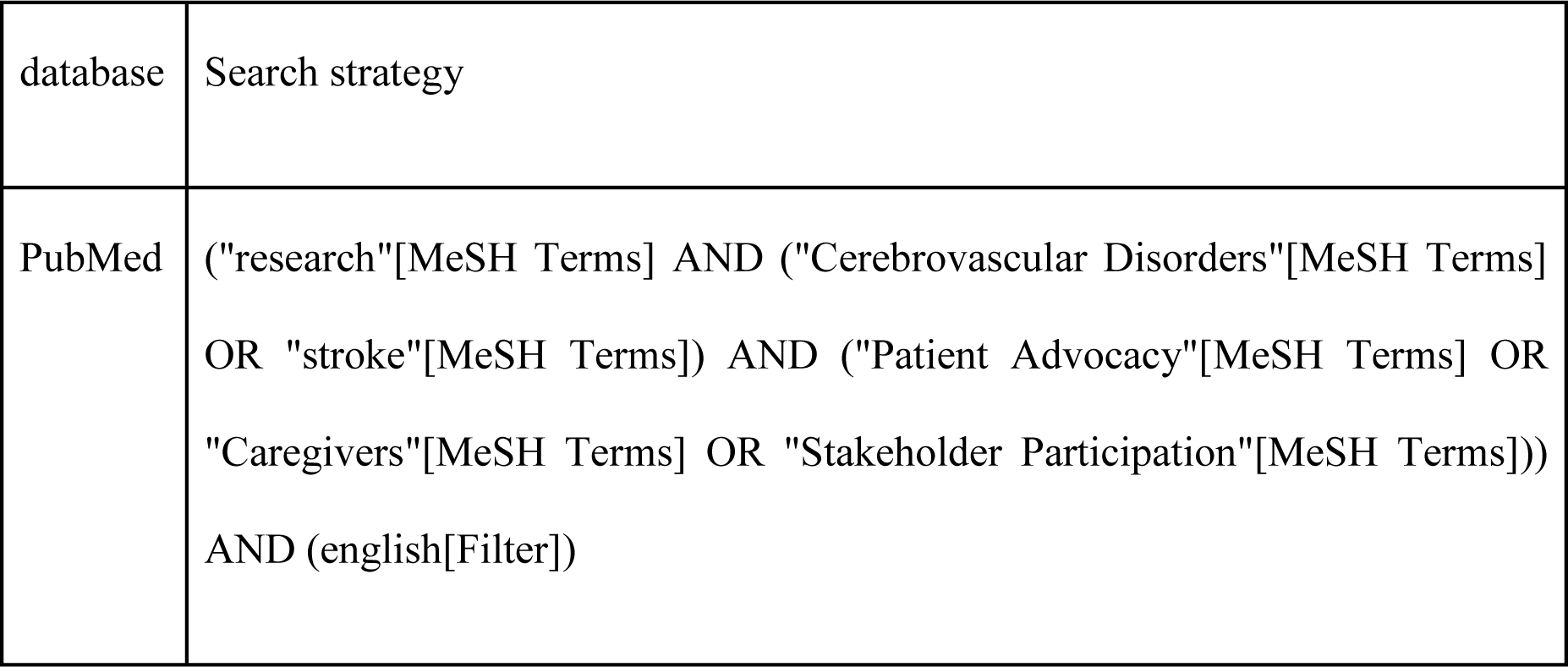
Search strategy of PubMed database.

## Supplemental file

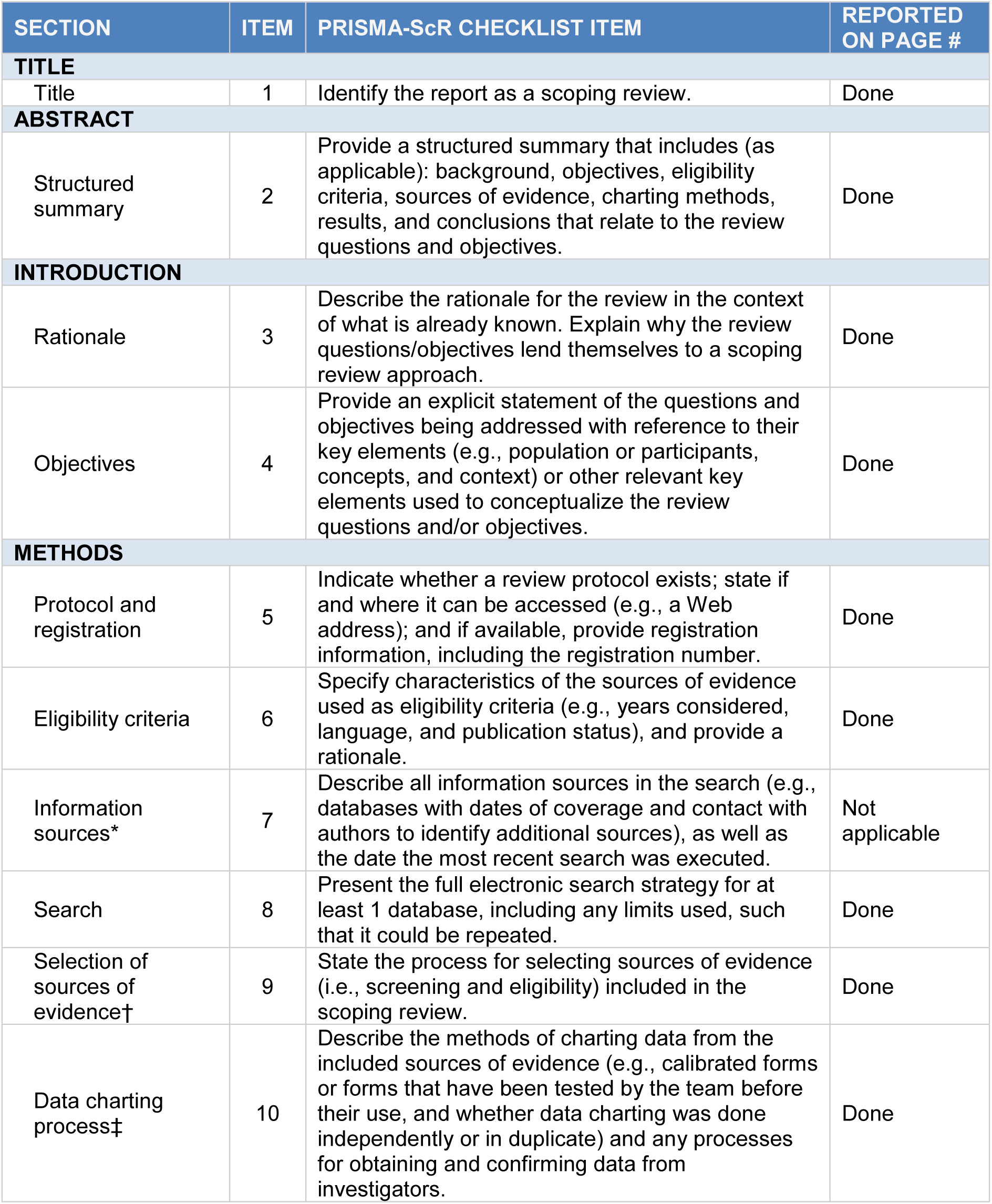
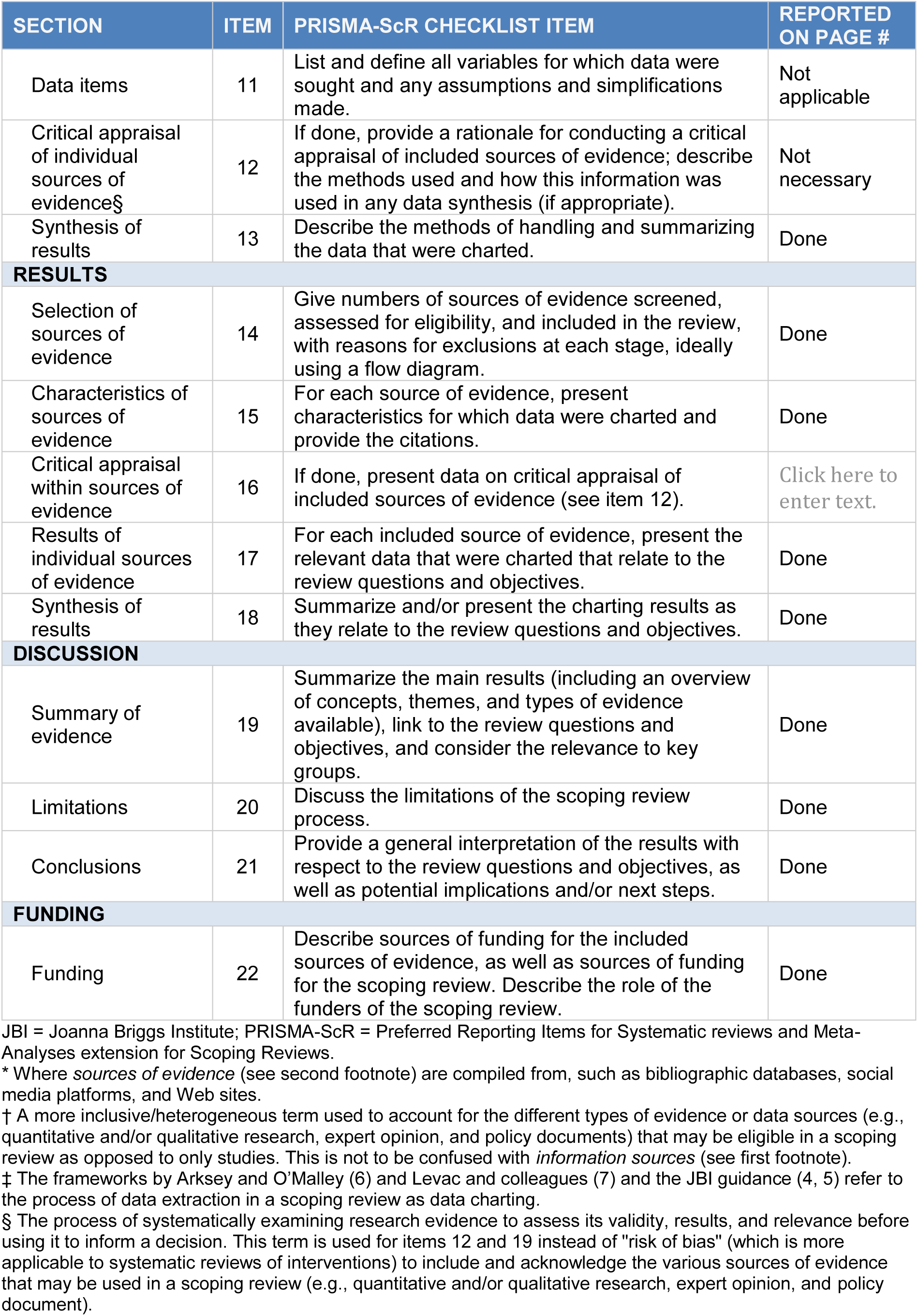
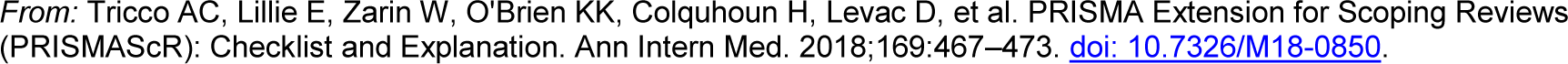
Preferred Reporting Items for Systematic reviews and Meta-Analyses extension for Scoping Reviews (PRISMA-ScR) Checklist.

